# Sleep Staging Foundation Models Encode Neural Disorder-Related EEG Representations that Generalize to Wakefulness

**DOI:** 10.1101/2025.11.15.25340316

**Authors:** Xavier A. Velez, Qichen Li, Samaneh Nasiri, Yashar Kiarashi, Gari D. Clifford

## Abstract

**Objective:** To leverage sleep foundation models trained on large datasets of polysomnography for neurological disorder detection during an awake state.

**Methods:** Three public resting-state electroencephalography (EEG) datasets from cognitively normal participants and patients with Alzheimer’s disease (AD), frontotemporal dementia (FTD), schizophrenia (SZ), and depression (MDD) were used. Embeddings from three pre-trained sleep staging foundation models were extracted and compared against a spectral model for AD, FTD, SZ, and MDD detection. Analyses used a subset of the available EEG channels, enabling comparisons between each EEG channel and the detection capability of the models.

**Results:** AD, FTD, and SZ classifications were best performed by the spectral model (AUROC=0.931, 0.878, 0.964), followed closely by SleepFM (AUROC=0.889, 0.863, 0.821), which was consistently the top-performing sleep staging model for wakeful disorder detection. All classifiers struggled to detect MDD and distinguish between AD and FTD (AUROC <0.80). For MDD classification, SleepFM (AUROC=0.737) outperformed the spectral model (AUROC=0.710). Between the top classifiers, the T4 and O2 channels were selected as part of the most informative group of electrodes for two-thirds of the disorder classification tasks.

**Conclusion:** The embeddings obtained from the sleep-staging foundation models captured meaningful information for detecting neurological disorders, even when using EEG from wakeful states. AD and FTD pathologies were the most sensitive to sleep staging analyses.

**Significance:** Our findings suggest that there is information in wakeful states proximal to sleep that reveals disease-induced neurological dysfunction, which can be represented by sleep staging models with no prior understanding of existing disorders.

## I. Introduction

Sleep clinics in the United States collect approximately 1.2 million polysomnographs (PSG) every year [1]. The abundance of PSG data facilitate the training of large-scale machine learning models [2] that can leverage the quantity of sleep data from established aggregated sources [3]–[8] to learn meaningful representations of sleep. Wakeful electroencephalography (EEG) is primarily collected in research studies or diagnostics of epilepsy or stroke, with an average recording duration of 20-90 minutes [9], compared to 5.8 hours as the average duration of PSG [10]. The availability of PSG underscores the effectiveness of deep learning models that evaluate sleep performance, and motivates the transition from traditional EEG to PSG, even for tasks involving wakeful brain states. In this work, we explore whether sleep representations extracted from models trained exclusively on PSG can be used to detect neurodegenerative and psychiatric disorders, promoting model generalization through data variability.

Alzheimer’s Disease (AD) and frontotemporal dementia (FTD) are forms of dementia that induce mood fluctuations, language difficulties, personality changes, and impairment of executive functions. AD is primarily associated with memory loss and presents in the hippocampus and neocortex, while FTD originates in the frontal and temporal lobes and is linked to behavioral changes. Both are frequently misdiagnosed: a 2013 study reported that 12–23% of AD diagnoses were not confirmed postmortem [11], and a 2017 paper found 24% of 916 AD patients received an incorrect diagnosis [12]. AD is characterized by amyloid beta buildup, a peptide normally cleared via glymphatic flow [13]. Slow-wave, or NREM sleep, accelerates this flow [14], revealing a cyclic relationship between diminishing sleep quality and disease progression [15], [16]. Although FTD lacks a distinct biomarker like amyloid beta, it is strongly associated with sleep disturbances, which can be more pronounced and more variable than in AD [17]. These findings motivate studying sleep-related changes as a method for better detecting and characterizing both dementias.

In this work, we extend the motivation for contextualizing neurodegenerative diseases through sleep to psychiatric disorders—specifically schizophrenia (SZ) and major depressive disorder (MDD). Neurodegenerative diseases have defined biomarkers and documented relationships with sleep, while psychiatric conditions, which are less understood, may also exhibit quantifiable sleep-linked physiological signatures. SZ research has progressed rapidly, with a 2022 study identifying variations in essential polysaturated fatty acids and lipid peroxidation metabolites [18], yet no biomarker is clinically established. MDD similarly lacks a clear biomarker, although cortisol has long been associated with MDD risk [19]. The DSM-5 [20] defines depression as persistent sadness or loss of interest, often accompanied by sleep disturbances, appetite changes, and fatigue, yet physiological biomarkers have not reliably characterized the disorder [21]. These gaps highlight continued need for improved physiological understanding of SZ and MDD.

Both psychiatric disorders are linked to changes in sleep. SZ patients frequently exhibit insomnia, reduced total sleep time, increased sleep onset latency, and reductions in slow-wave sleep [22], with increased prevalence of obstructive sleep apnea and circadian disorders [23]. MDD similarly shows disrupted sleep quality and duration, forming a cyclic relationship in which poor sleep both results from and worsens the disorder through emotional dysregulation [24]. Insomnia accounts for 88% of sleep disorders comorbid with MDD [25]. These strong links justify exploring sleep-related EEG features as markers for both psychiatric and neurodegenerative conditions.

EEG has historically been used to identify dementia and psychiatric disorders. On-scalp EEG captures electrical potentials linked to neural activity in shallow brain regions, enabling correlations between brain activity and neurological conditions. Although consistent and generalizable EEG biomarkers have not yet been clinically established, EEG remains useful for detecting AD, FTD, SZ, and MDD. Traditional machine learning models often rely on a single dataset, achieving favorable results but limiting generalizability to new cohorts or conditions. In this work, we leverage pre-trained sleep staging foundation models trained on PSG datasets to extract information relevant to neurological conditions. These sleep models are applied to resting-state, eyes-closed EEG, based on the hypothesis that sleep foundation models inherently encode neurological disorder–related features and that these learned representations can be exploited during wakefulness. This transfer learning framework may reveal a network capable of detecting and distinguishing multiple neurological conditions from wakeful EEG using sleep-guided embeddings.

## II. Related Works

Numerous computational works have approached AD, FTD, SZ, and MDD identification. However, few have applied the same machine learning process to various datasets across multiple conditions. While dementia is symptomatically different from psychiatric disorders, there may be elements of network dysfunction that a transfer learning model can extract and leverage to classify each condition, providing a more comprehensive and concise detection process than existing classification paradigms. Regardless, studies that independently classify AD, FTD, SZ, and MDD offer baselines for model performance with comparable experimental designs.

### A. Alzheimer’s Disease & Frontotemporal Dementia

Previous studies have applied AD and FTD detection using the same dataset described in the Dataset section. The initial publication by Miltiadous et al. [26] introduced an approach comparing classical machine learning methods based on relative bandpower features from delta, theta, alpha, beta, and gamma bands. Models including LightGBM [27], SVM [28], kNN [29], MLP [30], and Random Forests [31] were trained for binary classifications between AD-CN and FTD-CN groups. A Zheng et al. [32] expanded on this dataset by introducing multi-threshold recurrence rate plots (MTRRP) to detect neurophysiological patterns. MTRRP features were used within an SVM classifier for multiple binary and multiclass tasks, demonstrating notable performance improvements over the earlier baseline results. Ma et al. [33] proposed defining mutual information between electrode pairs as the primary features for classification. This approach used SVMs to perform AD-CN, FTD-CN, and AD-FTD classifications, achieving competitive or superior performance across most comparisons.

These studies demonstrate that AD and FTD can be effectively detected and distinguished using this dataset. The later works also identified relevant spatial information from different brain regions, providing context for distinguishing AD, FTD, and CN groups. A review by Nardone et al. [34] highlighted correlations between cortical oscillatory activity and dementia severity but emphasized the need for biomarkers capable of distinguishing individual patients. This demonstrates the importance of EEG representations that generalize beyond the conditions and data used during training. The discussed models were all trained and tested on the same dataset, potentially limiting generalizability, and motivating the need for approaches using learned parameters from external data sources.

### B. Schizophrenia

The public SZ dataset used in this work (of *N* = 28 individuals) has been tested in several other publications [35], [36]. Bagherzadeh et al. [35] tested an ensemble of large convolutional networks with partial directed coherence, direct directed transfer function, and transfer entropy across sequential time windows to detect SZxCN. The paper reports near perfect results. This is a common trend seen when using this dataset, potentially suggesting overfitting, which would limit the model’s ability to generalize to unseen participants. Aich et al. [36] used this dataset to test their approach of image encoding and wrapper-based feature selection using pre-trained deep models with subsequent K-NN clustering for final classification. Similar to the findings of Bagherzadeh et al., near-perfect accuracies were reported using this dataset. The two works present a pattern of this smaller dataset being shown as fully separable, likely as a result of overfitting. It is important to note that the intent of this work is not necessarily to compete in terms of high accuracies, especially on a public dataset that has been shown to have almost perfectly separable classes. Instead, we aim to explore whether the sleep staging foundation models will pick up on this separability, which, based on the accuracy, can be regarded as non-generalizable, or whether they may find a different mode of separation that might suggest a more balanced approach to SZ detection at the cost of performance.

### C. Depression

Depression detection using EEG has been frequently studied in the last decade, with convolutional layers leading the way for feature extraction methods as deep learning continues to be established as a favorable approach in EEG analysis [37]. Studies using the event-related potentials from the Multimodal Open Dataset for Mental Disorder Analysis (MODMA) depression dataset [38] have used CNN-based models [39] and EEGNet, a compact CNN for brain–computer interfaces [40] [41]. This portion of the dataset uses a dot-probe task with three kinds of emotional-neutral face pairs as the task stimuli. This approach requires participants to press a button when identifying an emotional-neutral face pair. However, a 2015 study found that patients with behavioral variant FTD had significant emotion recognition deficits when compared to an AD and CN group [42]. A dot-probe task that relies on emotion recognition queues from the brain may improve depression detection accuracies in otherwise healthy cohorts, but may not produce reliable results for patients with FTD or other disorders that impair emotion recognition functions.

To ensure compatibility with the relevant population, resting-state tasks have been chosen for comparison to this work, despite resulting in lower accuracies than other methods. The same MODMA dataset contains resting-state EEG recordings, which have been used in prior works employing deep learning approaches [43], [44]. While there are some works that reliably partition this dataset, classical machine learning approaches often perform much poorer in comparison to the deep learning models for this dataset [45]. Regardless, the prior works using this dataset will enable reasonable comparison to the methods proposed in this work.

## III. Datasets

### A. AD and FTD Dataset

The AD and FTD dataset applied in this work originates Miltiadous et al. [46]. Table I shows the participant demographics and Mini-Mental State Examination (MMSE) [47] for each group. A total of 88 participants (Female=44) were recruited. Out of these participants, 36 were diagnosed with AD, 23 were diagnosed with FTD, and 29 were CN. The Diagnostic and Statistical Manual of Mental Disorders, 3rd ed., revised (DSM-IIIR, DSM IV, ICD-10) [48] and the National Institute of Neurological, Communicative Disorders and Stroke—Alzheimer’s Disease and Related Disorders Association (NINCDS—ADRDA) [49] were used for diagnostics, and patients in the AD group with dementia comorbidities were excluded. For participants with AD or FTD, the median time since diagnosis was 25 months, with an interquartile range spanning 24-28.5 months. The MMSE was also administered during the intake session, providing a standard indicator of cognitive decline or impairment. The sex distribution between the groups was evaluated using a chi-squared test of independence and found statistically significant differences between the three groups (sex *X*^2^ = 6.78, *p* = 0.034). Age and MMSE differences were analyzed using a one-way ANOVA, finding non-significant differences in ages between the groups (*F* = 2.22, *p* = 0.115) and significant differences in MMSE scores (*F* = 119.74, *p* = 1.89e-25). While there were sex differences of note, the MMSE scores demonstrate an expected variation in cognitive function between groups.

**TABLE I:** Participant Demographics by Group. *±* indicates the standard deviation of the measured variable. Conventionally, MMSE scores below 24 are used as indicators of cognitive impairment.

|  | AD | FTD | CN |
| --- | --- | --- | --- |
| Female/Male | 24/12 | 9/14 | 11/18 |
| Age (Years) | 63.4 ± 7.8 | 63.7 ± 8.0 | 67.9 ± 5.3 |
| MMSE | 17.8 ± 4.4 | 22.2 ± 2.6 | 30.0 ± 0 |

EEG data was recorded from all 88 participants using a Nihon Kohden EEG 2100, a device commonly used in clinical settings. A total of 19 on-scalp electrodes were placed according to the 10-20 international system with a sampling rate of 500 Hz. Two reference electrodes on the mastoids were used to ensure skin impedance was below 5*k*Ω. Electrode Cz was the common reference in the referential montage included. A 500 Hz sampling rate was used, with the following amplifier parameters: sensitivity = 10uV/mm, time constant = 0.3s, and the high-frequency filter cutoff = 70 Hz. Additional recording information can be found in [26].

All data collected was done in a resting state eyes closed task in which the participants sat still upright. Each participant underwent a single session. A total of 485.5 minutes of EEG data for the AD group (average of 13.9 *±* 2.4 minutes), 276.5 minutes for the FTD group (average of 12.3 *±* 2.2 minutes), and 402 minutes for the CN group (average of 14.0 *±* 1.2 minutes). The second-to-eighth minutes (total seven minutes) of recordings were selected from each participant to standardize the length of time across groups and individuals in this dataset.

### B. Schizophrenia Dataset

The SZ dataset applied in this work was collected by researchers at the Polish Academy of Sciences [50]. Table II shows the participant demographics for each group. In this dataset, 28 participants were included, with balanced classes of 14 for both SZ and CN. All subjects included in the SZ class met international standards for paranoid schizophrenia diagnoses (see original publication for details on inclusion and exclusion criteria). Participants in the control group were recruited to match the age and sex of the SZ group, leading to non-significant differences in either demographic. A total of 15 minutes of 19-channel scalp EEG following the 10-20 system were recorded for each participant. However, to match the preprocessing steps for the AD and FTD dataset, only the second to eighth minute were selected.

**TABLE II:** Schizophrenia Dataset Demographics by Group. *±* indicates the standard deviation of the measured variable.

|  | SZ | CN |
| --- | --- | --- |
| Female/Male | 7/7 | 7/7 |
| Age (Years) | 28.1 ± 3.7 | 27.8 ± 3.2 |

### C. Depression Dataset

The MDD dataset applied in this work was collected by researchers at Lanzhou University [38], with a total of 53 subject (24 MDD, 29 CN). Table III shows the participant demographics and Pittsburgh Sleep Quality Index (PSQI) for each group. Several other quality of life and depression screening assessments were administered to all participants. However, given the relevance of sleep in this work, only the PSQI is reported in Table III, while all other metrics can be found in the original publication. The sex distribution between the two groups was evaluated using a chi-squared test of independence and found no statistically significant differences (sex X2 = 0.675, p = 0.411). Age, education, and PSQI significance between groups were analyzed using one-way ANOVAs, finding non-significant differences in ages between the groups (F = 0.046, p = 0.832), and significant differences in years of education (F = 10.3, p = 0.002) and PQSI (F = 87.9, p = 1.12e-12). While the statistically significant differences in reported sleep quality provide a foundation for the application of sleep staging models on this dataset, the difference in years of education presents an additional covariate not controlled for in the analyses of this work.

**TABLE III:** Major Depressive Disorder Dataset Demographics by Group. *±* indicates the standard deviation of the measured variable.

|  | MDD | CN |
| --- | --- | --- |
| Female/Male | 11/13 | 9/20 |
| Age (Years) | $30.9 \pm 10.2$ | $31.4 \pm 9.0$ |
| Education (Years) | $13.3 \pm 3.7$ | $16.0 \pm 2.3$ |
| PSQI | $12.5 \pm 4.1$ | $4.1 \pm 2.1$ |

**TABLE IV:** ADxCN Classification Performance. The table below shows the AUROC, AUPRC, sensitivity, and specificity for each of the final models tested on the ADxCN classification task.

| Features | AUROC | AUPRC | Sens | Spec |
| --- | --- | --- | --- | --- |
| Alpha Power | 0.931±0.08 | 0.867±0.21 | 0.929±0.14 | 0.933±0.12 |
| SleepFM Emb. | 0.886±0.13 | 0.813±0.21 | 0.846±0.18 | 0.926±0.10 |
| Single ST N1 Emb. | 0.602±0.23 | 0.669±0.25 | 0.567±0.32 | 0.637±0.23 |
| Multi ST N2 Emb. | 0.746±0.15 | 0.686±0.25 | 0.788±0.21 | 0.705±0.21 |

**TABLE V:**
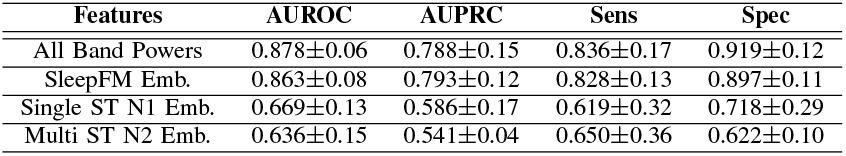
FTDxCN Classification Performance. The table below shows the AUROC, AUPRC, sensitivity, and specificity for each of the final models tested on the FTDxCN classification task.

| Features | AUROC | AUPRC | Sens | Spec |
| --- | --- | --- | --- | --- |
| All Band Powers | 0.878±0.06 | 0.788±0.15 | 0.836±0.17 | 0.919±0.12 |
| SleepFM Emb. | 0.863±0.08 | 0.793±0.12 | 0.828±0.13 | 0.897±0.11 |
| Single ST N1 Emb. | 0.669±0.13 | 0.586±0.17 | 0.619±0.32 | 0.718±0.29 |
| Multi ST N2 Emb. | 0.636±0.15 | 0.541±0.04 | 0.650±0.36 | 0.622±0.10 |

**TABLE VI:** ADxFTD Classification Performance. The table below shows the AUROC, AUPRC, sensitivity, and specificity for each of the final models tested on the ADxFTD classification task.

| Features | AUROC | AUPRC | Sens | Spec |
| --- | --- | --- | --- | --- |
| Theta + Beta Power | $0.775 \pm 0.09$ | $0.778 \pm 0.14$ | $0.852 \pm 0.14$ | $0.698 \pm 0.25$ |
| SleepFM Emb. | $0.694 \pm 0.16$ | $0.733 \pm 0.16$ | $0.819 \pm 0.19$ | $0.569 \pm 0.35$ |
| Single ST Wake Emb. | $0.621 \pm 0.15$ | $0.709 \pm 0.05$ | $0.688 \pm 0.22$ | $0.555 \pm 0.18$ |
| Multi ST All Emb. | $0.563 \pm 0.16$ | $0.645 \pm 0.18$ | $0.783 \pm 0.15$ | $0.343 \pm 0.24$ |

**TABLE VII:**
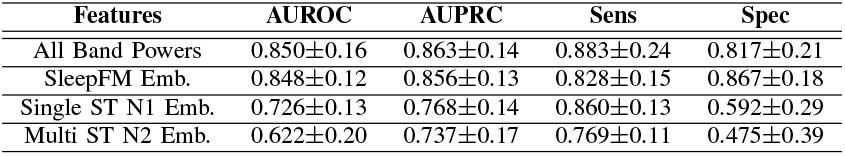
AD+FTDxCN Classification Performance. The table below shows the AUROC, AUPRC, sensitivity, and specificity for each of the final models tested on the AD+FTDxCN classification task.

| Features | AUROC | AUPRC | Sens | Spec |
| --- | --- | --- | --- | --- |
| All Band Powers | 0.850±0.16 | 0.863±0.14 | 0.883±0.24 | 0.817±0.21 |
| SleepFM Emb. | 0.848±0.12 | 0.856±0.13 | 0.828±0.15 | 0.867±0.18 |
| Single ST N1 Emb. | 0.726±0.13 | 0.768±0.14 | 0.860±0.13 | 0.592±0.29 |
| Multi ST N2 Emb. | 0.622±0.20 | 0.737±0.17 | 0.769±0.11 | 0.475±0.39 |

**TABLE VIII:**
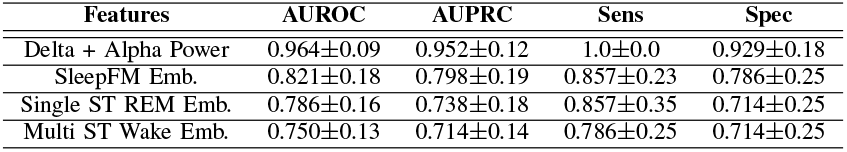
SZxCN Classification Performance. The table below shows the AUROC, AUPRC, sensitivity, and specificity for each of the final models tested on the SZxCN classification task.

Resting-state EEG was recorded using a 128-channel EEG. To keep as consistent as possible with the other datasets in this work, only the 19 channels associated with the 10-20 system were selected for preprocessing and analysis in this work, while all other channels were ignored. All recordings in this dataset were collected in a five-minute period, preventing the full seven-minute analyses applied to the other datasets. Instead, the second-to-fifth minutes of recordings were indexed for a total of four minutes worth of recording per participant. As alluded to in the related works, the MODMA dataset has a resting-state and task-based portion. Only the resting-state EEG was selected for this work.

## IV. Methods

While the primary aim of this work is to introduce a transfer learning approach to neurodegenerative and psychiatric disorder detection, a classical feature-based approach using EEG frequencies with XGBoost and SVM classifiers was first applied as a baseline, demonstrating an approach comparable to [26]. Once this baseline was established, a pretrained deep learning model was used to extract embeddings from the dataset. These were then tested on several classifiers in classical and transfer learning contexts.

### A. Preprocessing

All preprocessing was done in MATLAB R2021b using EEGLab version 2023.1 [51], primarily for the utilization of the artifact subspace reconstruction feature, which is a widely used standard for EEG preprocessing. The datasets were preprocessed using the following pipeline: 1) FIR Bandpass Butterworth Filer with pass band 0.5-45Hz, 2) signal rereferencing to the average of all nineteen electrodes, and 3) artifact subspace reconstruction, correcting artifacts that exceeded a maximum standard deviation of 17 within a 0.5 second window. The preprocessing steps applied in this work follow the specifications of the dementia dataset’s original pipeline [46] without the inclusion of independent component analysis (ICA) for eye or jaw artifact removal. However, given the importance of brain region identification in the interpretability of this work, it was decided to omit the ICA step. As a result, all data was re-preprocessed in EEGLab instead of using the original preprocessing while following the same guidelines. This process was applied to all three datasets used. For the MODMA dataset, 128 EEG channels were available. Regardless, the 19 channels described by the 10-20 system (excluding A1 and A2) were isolated from the array and preprocessed independently.

### B. Feature Extraction

#### 1) Spectral Features

Spectral features were first used to establish a baseline for binary classifications. Power spectral density (PSD; *NFFT* = 2^12^, *Overlap* = 2^8^, linear detrend) was computed independently for each channel and participant using a Dolph–Chebyshev window with an alpha attenuation parameter (*α* = 5) for 100 dB attenuation [52]. PSDs were separated into canonical frequency bands: Delta (0.5–4 Hz), Theta (4–8 Hz), Alpha (8–12 Hz), and Beta (16–25 Hz). Band powers including delta, theta, alpha, beta, all bands, and combinations of two were tested as the sole features across two classical machine learning models (Figure 1). For every test, five channels were selected under a channel sparsity constraint described in the supplementary information. These experiments aimed to determine whether spectral features (without temporal information) could adequately detect AD, FTD, SZ, or MDD, and to identify which features and electrodes were most informative for classification.

**Fig. 1:**
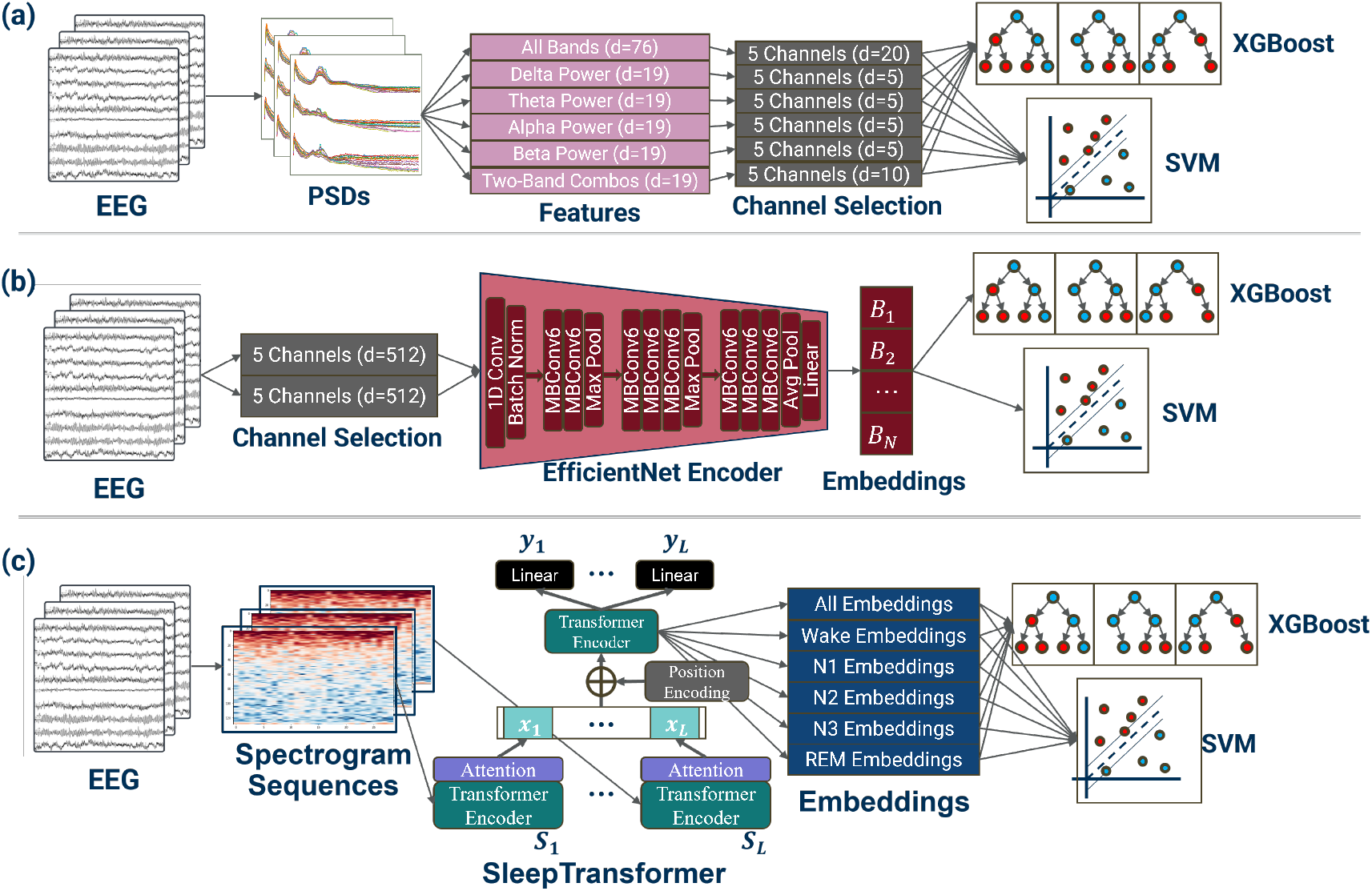
Overview of classification pipelines. Feature extraction and classification using spectral features (a), SleepFM embeddings (b), and the single- and multi-channel SleepTransformer embeddings (c). These pipelines were repeated for the following classification tasks: ADxCN, FTDxCN, ADxFTD, AD+FTDxCN, SZxCN, and MDDxCN.

#### 2) SleepFM Embeddings

While spectral features provided a clear baseline and useful context for electrode importance, we hypothesized that an embedding-based representation would capture richer information through transfer learning. We selected SleepFM-codebase [53], an earlier publicly released version of a larger model [54], trained on more than 100,000 hours of PSG data from over 14,000 participants. Its publicly available weights allowed us to leverage the full pretrained performance without retraining on our smaller dataset. The original model uses an EfficientNet [55] architecture to extract embeddings across multiple modalities (EEG, ECG, respiratory) and applies contrastive learning for future disease prediction. Although our dataset involves eyes-closed recordings rather than sleep, we hypothesized SleepFM would generalize. Because our data contained only EEG, we used only the pretrained EEG EfficientNet [53] to generate embeddings and did not use the contrastive learning component. To maintain compatibility with SleepFM, we matched its input/output structure: the model accepts exactly five EEG channels. To preserve channel interpretability, we chose electrode selection rather than dimensionality reduction (e.g., PCA); the same selection method used for the spectral features was applied here (details in the supplementary information). The dementia dataset was sampled at 500 Hz, and the schizophrenia and depression datasets (sampled at 250 Hz) were upsampled to 500 Hz via linear interpolation. The resulting SleepFM segment output 512-dimensional embeddings, which were used for AD, FTD, SZ, and MDD classification.

#### 3) SleepTransformer Embeddings

In addition to the original pre-trained SleepFM model, another deep sleep staging model, SleepTransformer [56], was applied to the described datasets. SleepTransformer was trained on the Sleep Heart Health Study (SHHS) dataset [8], and reported favorability for awake-state classification. Given the transfer learning approach of this work to resting-state eyes closed EEG data, a model that classifies wakefulness with high accuracy was found desirable. This model applies self-attention modules through transformer architectures at the epoch-level (30 seconds) and the sequence-level (21 epochs), producing partitioned embeddings for each relevant sleep stage (wake, N1, N2, N3, REM). The inputs to the model are 30-second spectrograms for each epoch. Raw, un-processed EEG signals were down-sampled to 100Hz and preprocessed using the pipeline applied in the original paper. Given that seven minutes of recorded EEG were extracted from the dementia and schizophrenia datasets, an overlap of 15 seconds was used to reach the recommended 21-epoch sequence length. The depression dataset, which we extracted four minutes of, required an overlap of 22 seconds to reach the desired sequence length. Once computed, the spectrograms were fed into the SleepTransformer, and embeddings were extracted. SleepTransformer was trained to take only one channel, and open weights were only available for the C4 electrode. Thus, this was the only channel selected and used to test this model, once again making use of the training the original authors performed.

#### 4) Multi-Channel SleepTransformer

Our group adapted the original SleepTransformer model [56] to use multiple channels, as opposed to a single channel. The multi-channel SleepTransformer extends the original single-channel architecture by enabling joint modeling of multiple EEG channels. To encode channel identity, a fixed sinusoidal channel embedding was added to the input feature representation before any attention operations. A dedicated channel-level Transformer encoder was then applied at the beginning of the network to explicitly model inter-channel dependencies through multihead self-attention. This module operates on the set of channels within each frame and produces channel-aware feature embeddings, which are subsequently reorganized and fed into the frame-level attention pooling mechanism. The resulting frame representations are processed by the sequence-level Transformer to capture long-range temporal structure across 121-epoch inputs.

For training, the multi-channel SleepTransformer was trained from scratch using computed spectrograms derived from full-night PSG recordings from 50 subjects randomly sampled across four major cohorts: SHHS [8], MROS [3], MGH [4], [5], and MESA [6]. The model was optimized end-to-end using a weighted focal loss to address class imbalance and enhance sensitivity to difficult-to-classify sleep stages.

Similar to the original SleepTransformer, this model takes in spectrogram epochs. Three-second epochs were used with a two-second overlap for one-second resolution within sequences. Data was down-sampled to 100Hz and pre-processed using the original pre-processing pipeline. The final multichannel SleepTransformer found channels C3, C4, F3, F4, O1, and O2 to be most informative for sleep staging. These were the final channels used for this work. For clarity throughout this work, the original version of the SleepTransformer will be referred to as the single-channel SleepTransformer, and the model trained by the authors of this work will be referred to as the multi-channel SleepTransformer. While different combinations of channels were not tested using these models, a feature of the single and multi-channel SleepTransformers includes partitions of embeddings for each possible stage of sleep. These embeddings were used together and separately to perform the classification task. This facilitates analysis of the sleep stage determined by each model to be most critical for disorder detection, even while the participants are awake.

### C. Classification and Channel Importance

We evaluated each feature set using eXtreme Gradient Boosting (XGBoost) [57] and Support Vector Machines (SVM) across several binary classification tasks. An 8-fold cross-validation scheme was used for hyperparameter selection and evaluation, with eight participants per fold for the dementia dataset, four per fold for the schizophrenia dataset, and six per fold for the depression dataset.

For the dementia dataset, ADxCN, FTDxCN, ADxFTD, and AD+FTDxCN were tested. Because AD and FTD originate from the same dataset, this setup enabled evaluation of each dementia type independently while also assessing each model’s ability to distinguish between them. In AD+FTDxCN, AD and FTD were grouped to represent generalized dementia-related neural dysfunction, providing context for whether models learned disease-specific signatures or broader dysfunction patterns. SZxCN and MDDxCN classifications were tested separately for the schizophrenia and depression datasets; because these datasets differ, SZ and MDD were not compared directly.

During channel selection for both spectral and SleepFM classifications, a running average of the area under the receiver operating characteristic curve (AUROC) was maintained for every valid channel combination. When a channel appeared in a given combination, the overall AUROC for that combination contributed to that channel’s average. This produced an average AUROC score for each of the 19 electrodes, which was then z-scored to compare regions. Because individual channels were never tested in isolation, these values should not be interpreted as independent channel importance. Instead, they represent each channel’s importance in combination with other selected electrodes, offering a more complete view of how surface regions interact to maximize disorder-related information.

## V. Results

### A. Alzheimer’s and Frontotemporal Dementia Classification Results

Figure 2*a* illustrates comparisons of AUROC for all of the previously described models using the dementia dataset.

**Fig. 2:**
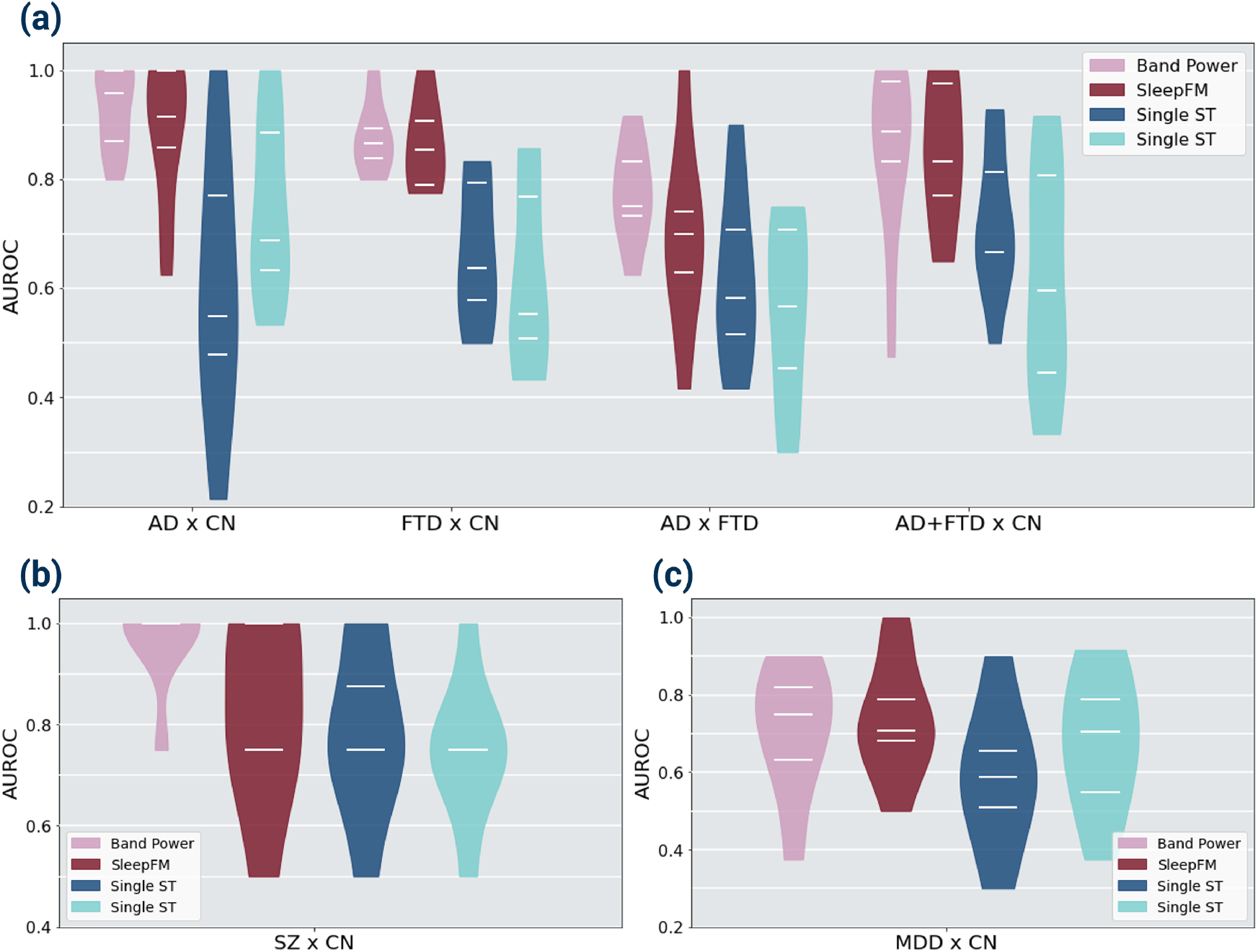
Classification Comparisons for All Tasks. Violin plots comparing AUROC distributions across folds for all classification tasks and models: (a) dementia dataset, (b) schizophrenia dataset, (c) major depressive disorder dataset. The three white lines shown in the violin plots represent the 25%, 50%, and 75% quantiles of the AUROC across folds.

ADxCN classification was best achieved by the spectral model using alpha power (*AUROC* = 0.931). SleepFM produced comparable results (*AUROC* = 0.886). The single-channel SleepTransformer (*AUROC* = 0.602) and multichannel SleepTransformer (*AUROC* = 0.746) performed poorly relative to both, using N1 and N2 embeddings, respectively. All 171 channel combinations were tested for the spectral and SleepFM models (Figure 3). For the spectral model, F7, Cz, T4, T5, and O2 were the most informative combination, with C4, T5, and O2 consistently important. SleepFM identified Fp2, F3, P3, T4, and O2 as the most informative, with F3, P3, P4, and O2 showing highest relative importance. Both models identified T4 and O2 as maximally important in combination with other electrodes. O2 specifically was shown to be the channel most indicative of AD for both models.

**Fig. 3:**
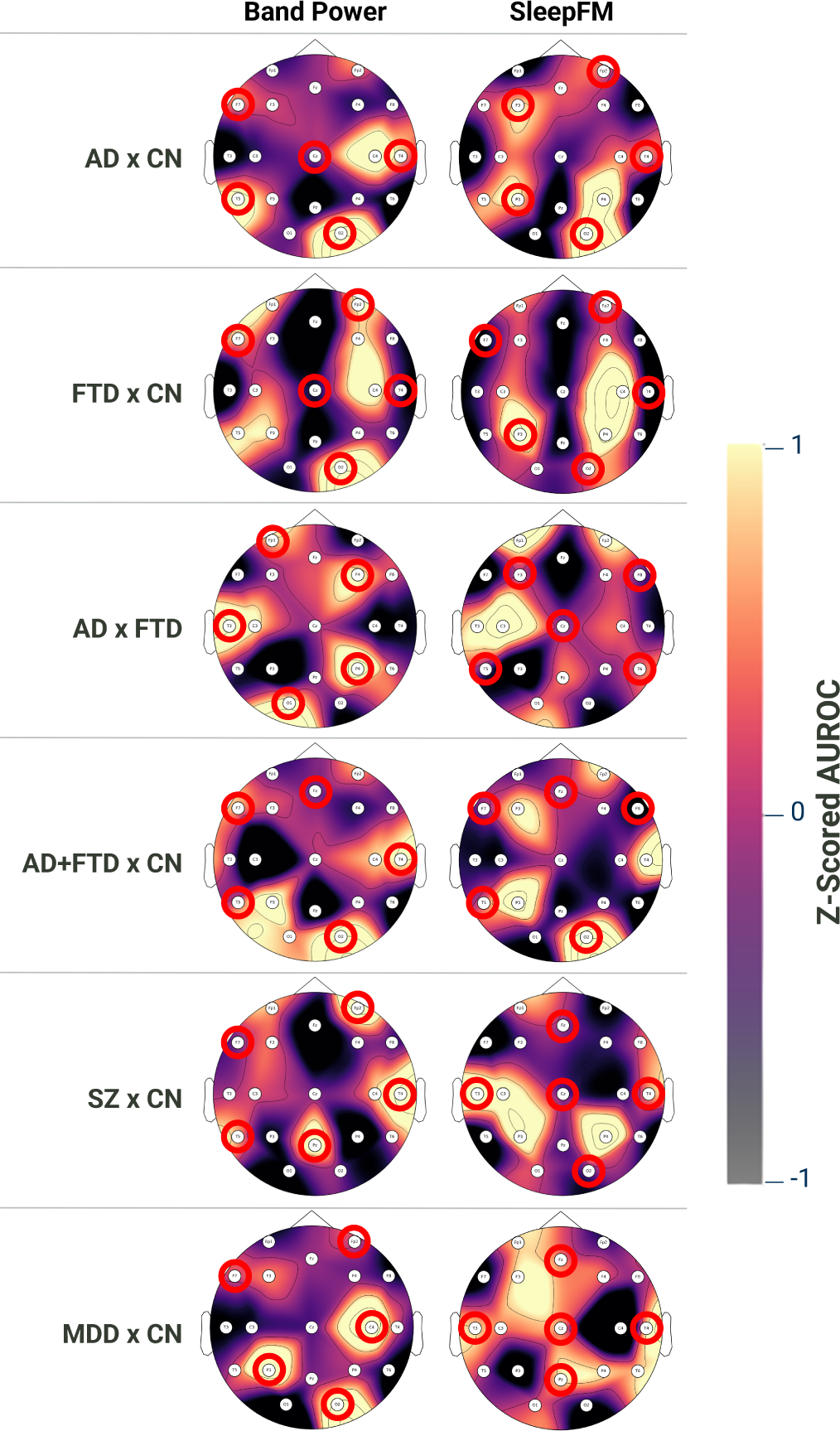
Electrode Saliency Plots for Spectral and SleepFM models. Channel maps follow the 10-20 EEG system. Light yellow areas represent surface regions with higher relative importance, while dark areas represent regions with lower relative importance. Electrodes circled in red belong to the optimally-chosen channel combination for the specified model.

For FTDxCN classification, the spectral model again performed best (*AUROC* = 0.878) using all band powers. SleepFM embeddings achieved similar accuracy (*AUROC* = 0.863). The single-channel SleepTransformer using N1 embeddings (*AUROC* = 0.669) and multi-channel SleepTransformer using N2 embeddings (*AUROC* = 0.636) performed poorly. Spectral model channel analyses for FTDxCN showed Fp2, F7, Cz, T4, and O2 as most informative. Additional nearby electrodes (Fp1, Fp2, F4, C4, P3, T5, O2) also contributed strongly. SleepFM found Fp2, F7, P3, T4, and O2 most informative. However, in the case of SleepFM, more of these channels are not part of the set of the most informative features across all combinations (C4, P3, P4). Both models showed symmetric importance across the axial midline, as seen in Figure 3.

After observing relatively high performance for AD and FTD classification against the cognitively normal cohort, a task distinguishing between the two forms of dementia would provide insight into whether the sleep staging models are encoding information related to disease-specific sleep-related perturbations, or neural dysfunction more broadly. For ADxFTD classification, the spectral model achieved the highest AUROC (0.775) using theta and beta power. SleepFM (0.694), single-channel SleepTransformer with wake embeddings (0.621), and multi-channel SleepTransformer with all embeddings (0.563) all performed worse. The spectral model identified Fp1, F4, P4, T3, and O1 as the most informative electrodes, which also overlapped with the most important across all combinations. SleepFM selected F3, F8, Cz, T5, and T8 as optimal, with Fp1, Fp2, C3, and T3 showing the most overall importance.

The final test for the dementia dataset involved combining the AD and FTD participants into one class for a broad dementia classification task. For AD+FTDxCN classification, the spectral model again performed best (*AUROC* = 0.850) using all band powers, with SleepFM performing similarly (*AUROC* = 0.848). Single- and multi-channel SleepTransformer models performed worse (*AUROC* = 0.756 and 0.622) using the N1 and N2 embeddings, respectively. Channel importance analysis (Figure 3) showed that the spectral model identified F7, Fz, T4, T5, and O2 as the most informative, with P3, T4, O1, and O2 most consistently important across channel combinations. SleepFM highlighted F3, P3, T4, and O2 across combinations, with its optimal combination being F7, F8, Fz, T5, and O2, closely matching the spectral model.

### B. Schizophrenia Classification Results

Figure 2b shows comparisons between all four models when classifying schizophrenia versus cognitively normal participants.

For schizophrenia detection, the spectral model achieved the highest AUROC (0.964) using alpha and delta power. SleepFM produced lower accuracy (*AUROC* = 0.821). The single-channel SleepTransformer (*AUROC* = 0.786) with REM embeddings, and the multi-channel SleepTransformer (*AUROC* = 0.750) with wake embeddings, were also outperformed by the spectral model. Electrode importance analysis for the spectral model identified Fp2, F7, Pz, T4, and T5 as the most informative channels, with Fp2, Pz, and T4 showing the highest overall importance. For SleepFM, the optimal combination was Fz, Cz, T3, T4, and O2, while C3, P3, P4, and T3 showed the greatest importance across combinations. These channel patterns differed substantially between the two models.

### C. Major Depressive Disorder Classification Results

Figure 2c shows comparisons between all four models when classifying major depressive disorder versus cognitively normal participants.

For MDDxCN classification, SleepFM achieved the highest AUROC (0.737). The spectral model followed (*AUROC* = 0.710) using delta and alpha power. The single-channel Sleep-Transformer (*AUROC* = 0.589) using wake embeddings, and the multi-channel SleepTransformer (*AUROC* = 0.680) using all embeddings, both performed below the spectral and SleepFM models. Electrode importance analysis for the spectral model identified Fp2, F7, C4, P3, and O2 as the optimal channel combination, with C4, P3, and O2 showing the highest relative importance across combinations. For SleepFM, the optimal combination was Fz, Cz, Pz, T3, and T4, with F3 and T4 showing the greatest overall importance. The spectral and SleepFM models exhibited contrasting importance patterns: C4, P3, and O2 were highly important in the spectral model but minimally important in SleepFM. Despite this, both models achieved similar performance levels (Table IX).

**TABLE IX:** MDDxCN Classification Performance. The table below shows the AUROC, AUPRC, sensitivity, and specificity for each of the final models tested on the MDDxCN classification task.

| Features | AUROC | AUPRC | Sens | Spec |
| --- | --- | --- | --- | --- |
| Delta + Alpha Power | 0.710±0.16 | 0.612±0.24 | 0.627±0.30 | 0.794±0.19 |
| SleepFM Emb. | 0.737±0.14 | 0.723±0.14 | 0.685±0.21 | 0.788±0.25 |
| Single ST Wake Emb. | 0.589±0.17 | 0.518±0.25 | 0.496±0.31 | 0.681±0.20 |
| Multi ST All Emb. | 0.680±0.17 | 0.609±0.18 | 0.581±0.27 | 0.779±0.16 |

## VI. Discussion

SleepFM embeddings produced performance close to the spectral model for ADxCN, indicating that learned sleep features can represent neural dysfunction during wakefulness. The poor results from both SleepTransformer variants suggest limited generalization for dementia detection when compared to spectral or SleepFM approaches. In this same task, the shared electrode importance patterns between the spectral and SleepFM models indicate that both methods emphasize temporo-occipital regions (particularly T4 and O2) for AD detection. Spectral and SleepFM models out-performed the original publication for ADxCN classification [26], which compared several classical models using relative EEG bandpower features (*Acc* = 0.770, *Sens* = 0.783, *Spec* = 0.809). The ADxCN classification was also higher than both related works [32], [33] (*Acc* = 0.877, *Sens* = 0.972, *Spec* = 0.759 and *Acc* = 0.769, *Sens* = 0.769, *Spec* = 0.720, respectively). These comparisons provide merit for the conclusions that electrode selectivity and sleep staging models can introduce representations of AD that effectively partition this dataset.

For FTDxCN classification, the comparable performance between the spectral and SleepFM models implies that SleepFM captures relevant disease-specific information. The symmetry of importance maps across hemispheres suggests that both models extract distributed cortical signals associated with FTD-related dysfunction. The original dataset publication in Miltiadous et al. (*Acc* = 0.731, *Sens* = 0.630, *Spec* = 0.786) and Ma et al., which used mutual information between electrodes (*Acc* = 0.904, *Sens* = 0.904, *Spec* = 0.879), both reported lower FTDxCN accuracies than the spectral and SleepFM models in this work. Zheng et al., however, outperformed all reported models (*Acc* = 0.827, *Sens* = 0.739, *Spec* = 0.897) for FTDxCN classification.

The ADxFTD results show reduced accuracy across all models in this work, indicating that sleep foundation models—and even spectral power features—struggle to distinguish between forms of dementia. The differing electrode selections between the spectral and SleepFM models further imply that SleepFM may encode more generalized markers of neural dysfunction related to sleep processes, rather than disease-specific signatures. While Miltiadous et al. did not test ADxFTD classification, Ma et al. (*Acc* = 0.915, *Sens* = 0.915, *Spec* = 0.867) and Zheng et al. (*Acc* = 0.729, *Sens* = 0.944, *Spec* = 0.391) reported higher accuracies than SleepFM. The spectral model in this work produced higher accuracies than the Zheng et al. classifier, but all accuracies were significantly lower than in Ma et al. for this task.

The AD+FTDxCN results reinforce the interpretation that SleepFM encodes more generalized features of neural dysfunction rather than disease-specific information. SleepFM matched spectral performance, suggesting that the model effectively encodes information associated with generalized dementia-related dysfunction. The overlap in informative channels, particularly in parietal and occipital areas, supports the conclusion that both models identify similar spatial patterns of neural disruption. However, the higher accuracies in separate ADxCN and FTDxCN tasks suggest that disease-specific features persist, preventing full generalization across dementia types. The primary aim of this classification task was to further contextualize the information encoded by the sleep staging foundation models. As such, the compared related works [26], [32], [33] did not perform this classification task.

The schizophrenia classification results followed the general trend observed in the dementia tasks, with the spectral model outperforming all others. However, the large performance gap between the spectral model and SleepFM suggests that the spectral model may be overfitting, given the limited dataset size (*n* = 28). This interpretation aligns with prior reports of similarly high spectral performance in Bagherzadeh et al. (*AUROC* = 0.998, *Sens* = 0.995, *Spec* = 1.0) [35] and Aich et al. (*Acc* = 0.9997, *Sens* = 0.9996, *Prec* = 0.9995) [36]. Further evidence of the ease of separability of this dataset is shown in the supplementary information. This emphasizes the need for validation on larger datasets to assess generalizability. Despite lower accuracy, the relatively strong SleepFM performance indicates that it encoded schizophrenia-related information. However, the distinct electrode importance patterns between the spectral and SleepFM models suggest they capture different underlying neural features, unlike the overlap observed in the dementia classification tasks.

Unlike all other classification tasks, the spectral model did not outperform the sleep staging models for MDDxCN, with SleepFM achieving the highest accuracy. This result suggests that the SleepFM embeddings capture neural dynamics relevant to MDD that may not be reflected in absolute band power features. The consistently poor performance of the SleepTransformer models across tasks indicates that not all sleep staging models encode equivalent levels of neural dysfunction-related information. The similar accuracies between the spectral and SleepFM models, despite their distinct electrode importance patterns, suggest that both struggle to fully characterize MDD, which aligns with the lower overall accuracies relative to other disorders. While these findings imply a possible link between MDD and sleep-related neural representations that generalize to wakefulness, they also highlight limitations in the SleepFM and SleepTransformer models’ ability to capture MDD-specific dysfunction during wakeful states. Prior works employing deep learning methods such as Wang et al. [43] and Liu et al. [44] reported much higher accuracy of 95% and 98%, respectively, using the MODMA dataset. While classical approches such as Sun et al. [45] reported lower accuracies (82.3%), these works all surpass the performance of all models in this work. The performance of SleepFM over the spectral model and variations in the channel importance demonstrate varying information between the models may inform a further understanding of the links between MDD and sleep. However, the use of the tested sleep staging models for MDDxCN classification is not justified when compared to related works.

In all classification tasks, the single- and multi-channel SleepTransformers produced lower accuracies than the spectral and SleepFM models. Despite these disparities, the reported results of these models show consistent disease-specific information, even if not at the magnitude of the other models. As discussed in the methods, these models were trained on limited amounts of data compared to SleepFM and were only tested using the channels defined during training. Our results demonstrate that these models can reliably produce strong results, even without copious amounts of data. This further solidifies the conclusion that some sleep staging models can inherently encode some elements of neural dysfunction regardless of the training size. However, the amount of training data and model architecture may have non-trivial implications on model performance.

The methods applied in this work are founded in the availability of large PSG datasets across multiple institutions and cohorts. While the scope of these techniques included leveraging pre-trained sleep staging models in a transfer learning approach, training deep learning architectures only on the classified wake states of the PSG may offer a more robust transfer of information to wakeful EEG.

## VII. Conclusion

Alzheimer’s disease, frontotemporal dementia, schizophrenia, and depression all have well-established links to sleep, motivating continued exploration of sleep-related neural patterns for disease detection. This work showed that classical spectral band-power analyses and transfer learning using sleep-staging foundation models perform similarly for disorder classification during wakeful states. Across ADxCN (*AUROC* = 0.931 vs. 0.886), FTDxCN (0.878 vs. 0.863), ADxFTD (0.775 vs. 0.694), AD+FTDxCN (0.850 vs. 0.848), SZxCN (0.964 vs. 0.821), and MDDxCN (0.710 vs. 0.737), spectral and SleepFM models produced the highest results. The relatively strong performance of SleepFM indicates that deep learning sleep-staging models inherently encode information relevant to neurodegenerative and psychiatric disorders, further supported by non-trivial SleepTransformer performance. Overlapping channel importance highlights alignment between SleepFM and spectral approaches. Overall, findings suggest that sleep-staging models capture disorder-specific signals but more broadly detect neural dysfunction, with limited ability to separate clinically similar conditions. Applying pretrained sleep models to wakeful EEG appears effective for neurodegenerative disorders and may offer a less overfit approach for schizophrenia detection. This opens the potential for characterizing these conditions through a sleep-informed representation while benefiting from fast, single-session EEG acquisition.

## VIII. Acknowledgements

GC owns stock in and is an advisor to NextSense, Inc., which partially funded this research study. The terms of this arrangement have been reviewed and approved by Emory University in accordance with Emory University Policy 7.7, Policy for Investigators Holding a Financial Interest in Research. XV is partially funded by unrestricted funds from ARCS Foundation. GC, YK and XV acknowledge support from the Alzheimer’s Association, The Michael J. Fox Foundation for Parkinson’s Research, and CurePSP Sleep Contributions to Neurodegeneration Grant Program (SCN-25-1470707).

## Data Availability

All data produced are available online at: https://openneuro.org/datasets/ds004504/versions/1.0.7 https://doi.org/10.18150/repod.0107441 https://modma.lzu.edu.cn/data/application/#data_1

