## Supplementary Tables & Figures for "Sleep Staging Foundation Models Encode Neural Disorder-Related EEG Representations that Generalize to Wakefulness"

### Supplementary Information

TABLE I: **Electrode Combination Mapping.** The table below lists all 19 channels and the list of channels that are not allowed to be in the same combination. This drastically reduces the number of five-channel combinations that can be tested from 11628, which is not computationally practical, to 171. These 171 channel combinations are all tested for spectral feature models and SleepFM embedding models.

| Channel | Not allowed combinations |
|---|---|
| Fp1 | Fp2, F3, F7, Fz |
| Fp2 | Fp1, F4, F8, Fz |
| F3 | Fp1, F7, Fz, C3, T3 |
| F4 | Fp2, F8, Fz, C4, T4 |
| F7 | Fp1, F3, C3, T3 |
| F8 | Fp2, F4, C4, T4 |
| Fz | Fp1, Fp2, F3, F4 |
| C3 | F7, Cz, P3, T3 |
| C4 | F8, Cz, P4, T4 |
| Cz | P3, P4, C3, C4 |
| P3 | C3, Pz, T3, T5, O1 |
| P4 | C4, Pz, T4, T6, O2 |
| Pz | P3, P4, O1, O2 |
| T3 | F3, F7, C3, P3, T5 |
| T4 | F4, F8, C4, P4, T6 |
| T5 | C3, P3, T3, O1 |
| T6 | C4, P4, T4, O2 |
| O1 | P3, Pz, T5, O2 |
| O2 | P4, Pz, T6, O1 |

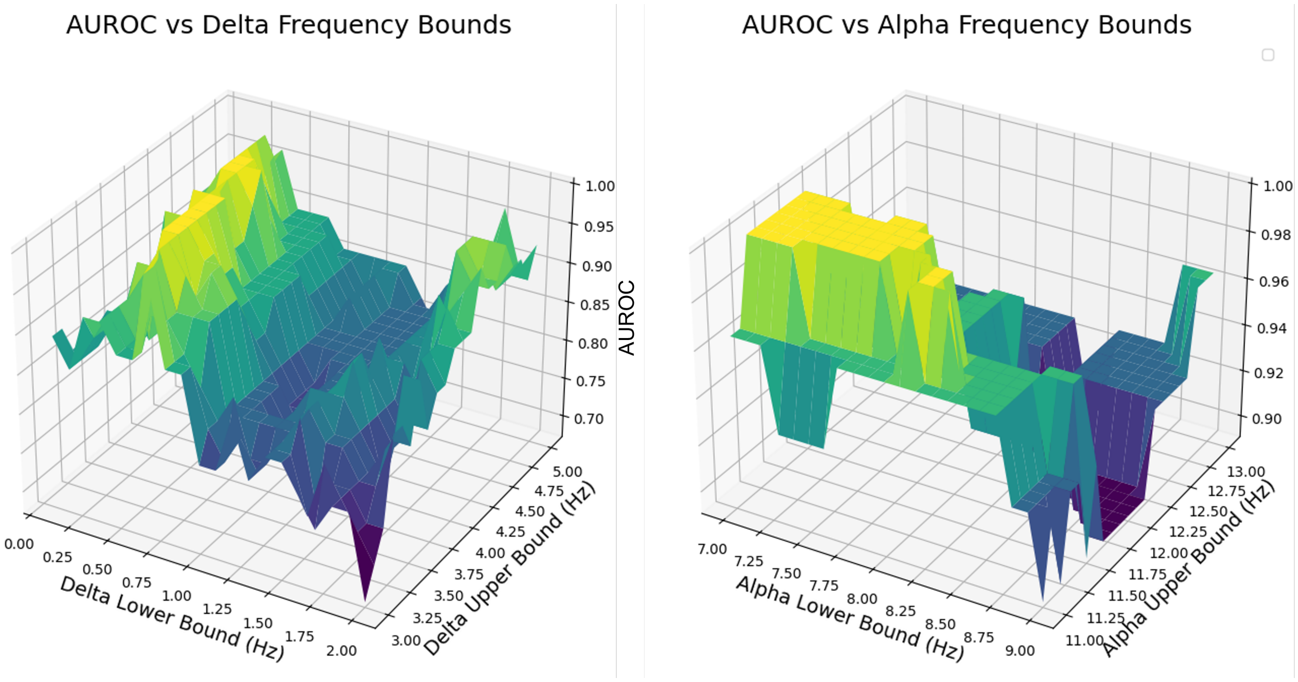


Fig. 1: **AUROC of SZxCN with Varying Bandpower Limits.** AUROC for SZxCN using grid search of bandpower limits in 0.25 Hz increments. Classification is performed using absolute band powers with both delta and alpha bands using channels Fp2, F7, Pz, T4, and T5. Variations in the delta band (alpha fixed to 7.5-11.5 Hz) shown on the left and variations in the alpha band (delta fixed to 0.5-4.0 Hz) shown on the right produce AUROC of 1.0 in various regions. This simple approach to classification demonstrates a simplicity to partitioning this dataset.
